# Weighted burden analysis in 200 000 exome-sequenced UK Biobank subjects characterises effects of rare genetic variants on BMI

**DOI:** 10.1101/2021.01.20.21250151

**Authors:** David Curtis

**Affiliations:** UCL Genetics Institute, UCL, Darwin Building, Gower Street, London WC1E 6BT; Centre for Psychiatry, Queen Mary University of London, Charterhouse Square, London EC1M 6BQ

**Keywords:** BMI, biobank, exome, *MC4R*, *PCSK1*, *SIRT1*, *ZBED6*, *NPC2*

## Abstract

**Introduction:** A number of genes have been identified in which rare variants can cause obesity. Here we analyse a sample of exome sequenced subjects from UK Biobank using BMI as a phenotype.

**Methods:** There were 199,807 exome sequenced subjects for whom BMI was recorded. Weighted burden analysis of rare, functional variants was carried out, incorporating population principal components and sex as covariates. For selected genes, additional analyses were carried out to clarify the contribution of different categories of variant. Statistical significance was summarised as the signed log 10 of the p value (SLP), given a positive sign if the weighted burden score was positively correlated with BMI.

**Results:** Two genes were exome-wide significant, *MC4R* (SLP = 15.79) and *PCSK1* (SLP = 6.61). In *MC4R*, disruptive variants were associated with an increase in BMI of 2.72 units and probably damaging nonsynonymous variants with an increase of 2.02 units. In *PCSK1*, disruptive variants were associated with a BMI increase of 2.29 and protein-altering variants with an increase of 0.34. Results for other genes were not formally significant after correction for multiple testing, although *SIRT1, ZBED6* and *NPC2* were noted to be of potential interest.

**Conclusion:** Because the UK Biobank consists of a self-selected sample of relatively healthy volunteers, the effect sizes noted may be underestimates. The results demonstrate the effects of very rare variants on BMI and suggest that other genes and variants will be definitively implicated when the sequence data for additional subjects becomes available.

This research has been conducted using the UK Biobank Resource.

## Introduction

Genome wide association studies (GWAS) detect large numbers of common variants showing statistically significant association with obesity although it can be difficult to interpret the biological processes underlying these signals (Müller et al., 2018). Additionally, a small number of genes have been identified in which very rare variants can have a major effect on body mass index (BMI) and their contribution and mechanisms have recently been reviewed (Thaker, 2017). In some of these, such as *LEP, LEPR, PCSK1* and *SIM1*, recessively acting variants cause deficiency of the gene product and this can result in obesity. In others, including *POMC* and *MC4R*, heterozygous variants have been reported to be causative. Dominantly and recessively acting *MC4R* variants together constitute the commonest causes of inherited early-onset obesity, with a prevalence of 0.5-0.6%. It is also recognised that other nonsynonymous variants in *MC4R* can be associated with lower BMI and can be protective against obesity (Stutzmann et al., 2007).

As sequence data becomes available for larger numbers of subjects, this makes it possible to explore the contribution of rare genetic variants to traits in the general population and we recently reported results obtained from analysing the association between rare variants and BMI in 50,000 exome-sequenced UK Biobank subjects (Curtis, 2021a). Although no gene was exome wide significant, the analysis did highlight some which were potentially of interest, including *LYPLAL1* and *NSDHL*. Since then, additional data has been released meaning that exome sequence data is now available for 200,000 of the 500,000 UK Biobank subjects (Szustakowski et al., 2020). Analyses of this larger dataset shows that it is better powered to detect rare variant effects and such analyses were successful in implicating, at exome-wide significance, genes previously recognised as risk factors for both hyperlipidaemia and type 2 diabetes (Curtis, 2021b, 2021c). Here, we apply the same approach as previously, using BMI as the phenotype in the enlarged sample.

## Methods

The UK Biobank dataset was downloaded along with the variant call files for 200,632 subjects who had undergone exome-sequencing and genotyping by the UK Biobank Exome Sequencing Consortium using the GRCh38 assembly with coverage 20X at 95.6% of sites on average (Szustakowski et al., 2020). UK Biobank had obtained ethics approval from the North West Multi-centre Research Ethics Committee which covers the UK (approval number: 11/NW/0382) and had obtained informed consent from all participants. The UK Biobank approved an application for use of the data (ID 51119) and ethics approval for the analyses was obtained from the UCL Research Ethics Committee (11527/001). All variants were annotated using the standard software packages VEP, PolyPhen and SIFT (Adzhubei et al., 2013; Kumar et al., 2009; McLaren et al., 2016). To obtain population principal components reflecting ancestry, version 2.0 of *plink* (https://www.cog-genomics.org/plink/2.0/) was run with the options *--maf 0*.*1 --pca 20 approx* (Chang et al., 2015; Galinsky et al., 2016). The phenotype was obtained from data field 21001-0.0, which records BMI at first assessment.

Using the same approach as described previously, the SCOREASSOC program was used to carry out a weighted burden analysis to test whether, in each gene, the weighted burden of sequence variants which were rarer and/or predicted to have more severe functional effects correlated with BMI (Curtis, 2021a). Attention was restricted to rare variants with minor allele frequency (MAF) <= 0.01. As previously described, variants were weighted by overall MAF so that variants with MAF=0.01 were given a weight of 1 while very rare variants with MAF close to zero were given a weight of 10 (Curtis, 2021a). Variants were also weighted according to their functional annotation using the GENEVARASSOC program, which was used to generate input files for weighted burden analysis by SCOREASSOC (Curtis, 2016, 2012). The weights were informed from the analysis of the effects of different categories of variant in *LDLR* on hyperlipidaemia risk (Curtis, 2021b). Variants predicted to cause complete loss of function (LOF) of the gene were assigned a weight of 100. Nonsynonymous variants were assigned a weight of 5 but if PolyPhen annotated them as possibly or probably damaging then 5 or 10 was added to this and if SIFT annotated them as deleterious then 20 was added. In order to allow exploration of the effects of different types of variant on disease risk the variants were also grouped into broader categories to be used in multivariate analyses as described below. The full set of weights and categories is displayed in Table 1. As described previously, the weight due to MAF and the weight due to functional annotation were multiplied together to provide an overall weight for each variant. Variants were excluded if there were more than 10% of genotypes missing or if the heterozygote count was smaller than both homozygote counts. If a subject was not genotyped for a variant then they were assigned the subject-wise average score for that variant. For each subject a gene-wise weighted burden score was derived as the sum of the variant-wise weights, each multiplied by the number of alleles of the variant which the given subject possessed. For variants on the X chromosome, hemizygous males were treated as homozygotes.

**Table 1.** The table shows the weight which was assigned to each type of variant as annotated by VEP, Polyphen and SIFT as well as the broad categories which were used for multivariate analyses of variant effects (Adzhubei et al., 2013; Kumar et al., 2009; McLaren et al., 2016).

| **VEP / SIFT / Polyphen annotation** | **Weight** | **Category** |
| --- | --- | --- |
| intergenic_variant | 0 | Unused |
| feature_truncation | 0 | Intronic, etc. |
| regulatory_region_variant | 0 | Intronic, etc. |
| feature_elongation | 0 | Intronic, etc. |
| regulatory_region_amplification | 1 | Intronic, etc. |
| regulatory_region_ablation | 1 | Intronic, etc. |
| TF_binding_site_variant | 1 | Intronic, etc. |
| TFBS_amplification | 1 | Intronic, etc. |
| TFBS_ablation | 1 | Intronic, etc. |
| downstream_gene_variant | 0 | Intronic, etc. |
| upstream_gene_variant | 0 | Intronic, etc. |
| non_coding_transcript_variant | 0 | Intronic, etc. |
| NMD_transcript_variant | 0 | Intronic, etc. |
| intron_variant | 0 | Intronic, etc. |
| non_coding_transcript_exon_variant | 0 | Intronic, etc. |
| 3_prime_UTR_variant | 1 | 3 prime UTR |
| 5_prime_UTR_variant | 1 | 5 prime UTR |
| mature_miRNA_variant | 5 | Unused |
| coding_sequence_variant | 0 | Unused |
| synonymous_variant | 0 | Synonymous |
| stop_retained_variant | 5 | Unused |
| incomplete_terminal_codon_variant | 5 | Unused |
| splice_region_variant | 1 | Splice region |
| protein_altering_variant | 5 | Protein altering |
| missense_variant | 5 | Protein altering |
| inframe_deletion | 10 | InDel, etc |
| inframe_insertion | 10 | InDel, etc |
| transcript_amplification | 10 | InDel, etc |
| start_lost | 10 | Unused |
| stop_lost | 10 | Unused |
| frameshift_variant | 100 | Disruptive |
| stop_gained | 100 | Disruptive |
| splice_donor_variant | 100 | Splice site variant |
| splice_acceptor_variant | 100 | Splice site variant |
| transcript_ablation | 100 | Disruptive |
| SIFT deleterious | 20 | Deleterious |
| PolyPhen possibly damaging | 5 | Possibly damaging |
| PolyPhen probably damaging | 10 | Probably damaging |

For each gene, multiple linear regression analysis was carried out including the first 20 population principal components and sex as covariates and a likelihood ratio test was performed comparing the likelihoods of the models with and without the gene-wise burden score. The statistical significance was summarised as a signed log p value (SLP), which is the log base 10 of the p value given a positive sign if the score is positively correlated with BMI.

Gene set analyses were carried out as before using the 1454 “all GO gene sets, gene symbols” pathways as listed in the file *c5*.*all*.*v5*.*0*.*symbols*.*gmt* downloaded from the Molecular Signatures Database at http://www.broadinstitute.org/gsea/msigdb/collections.jsp (Subramanian et al., 2005). For each set of genes, the natural logs of the gene-wise p values were summed according to Fisher’s method to produce a chi-squared statistic with degrees of freedom equal to twice the number of genes in the set. The p value associated with this chi-squared statistic was expressed as a minus log10 p (MLP) as a test of association of the set with BMI.

For selected genes, additional analyses were carried out to clarify the contribution of different categories of variant. As described previously, multiple linear regression analyses were performed on the counts of the separate categories of variant as listed in Table 1, again including principal components and sex as covariates, to estimate the effect size for each category (Curtis, 2021b). The mean effect on BMI for each category was estimated along with the standard error and the Wald statistic was used to obtain a p value. The associated p value was converted to an SLP, again with the sign being positive if the mean count was positively correlated with BMI. In these analyses, stop variants and frameshift variants were considered jointly as “disruptive variants” and splice site variants were considered separately, although all three types of variant might generally be expected to have a similar LOF effect.

Data manipulation and statistical analyses were performed using GENEVARASSOC, SCOREASSOC and R (R Core Team, 2014).

## Results

There were 199,807 exome sequenced subjects for whom BMI was recorded. There were 20,384 genes for which there were qualifying variants, meaning that the critical threshold for the absolute value of the SLP to declare a result as formally statistically significant is -log10(0.05/20384) = 5.61. This threshold was met by two genes, *MC4R* (SLP = 15.79) and *PCSK1* (SLP = 6.61). The quantile-quantile (QQ) plot for the SLPs obtained for all genes except *MCR4* is shown in Figure 1. This shows that the test appears to be well-behaved and conforms fairly well with the expected distribution. Omitting the genes with the 100 highest and 100 lowest SLPs, which might be capturing a real biological effect, the gradient for positive SLPs is 1.23 with intercept at -0.0005 and the gradient for negative SLPs is 1.03 with intercept at 0.02, indicating only moderate inflation of the test statistic for those genes showing a positive correlation.

**Figure 1.**
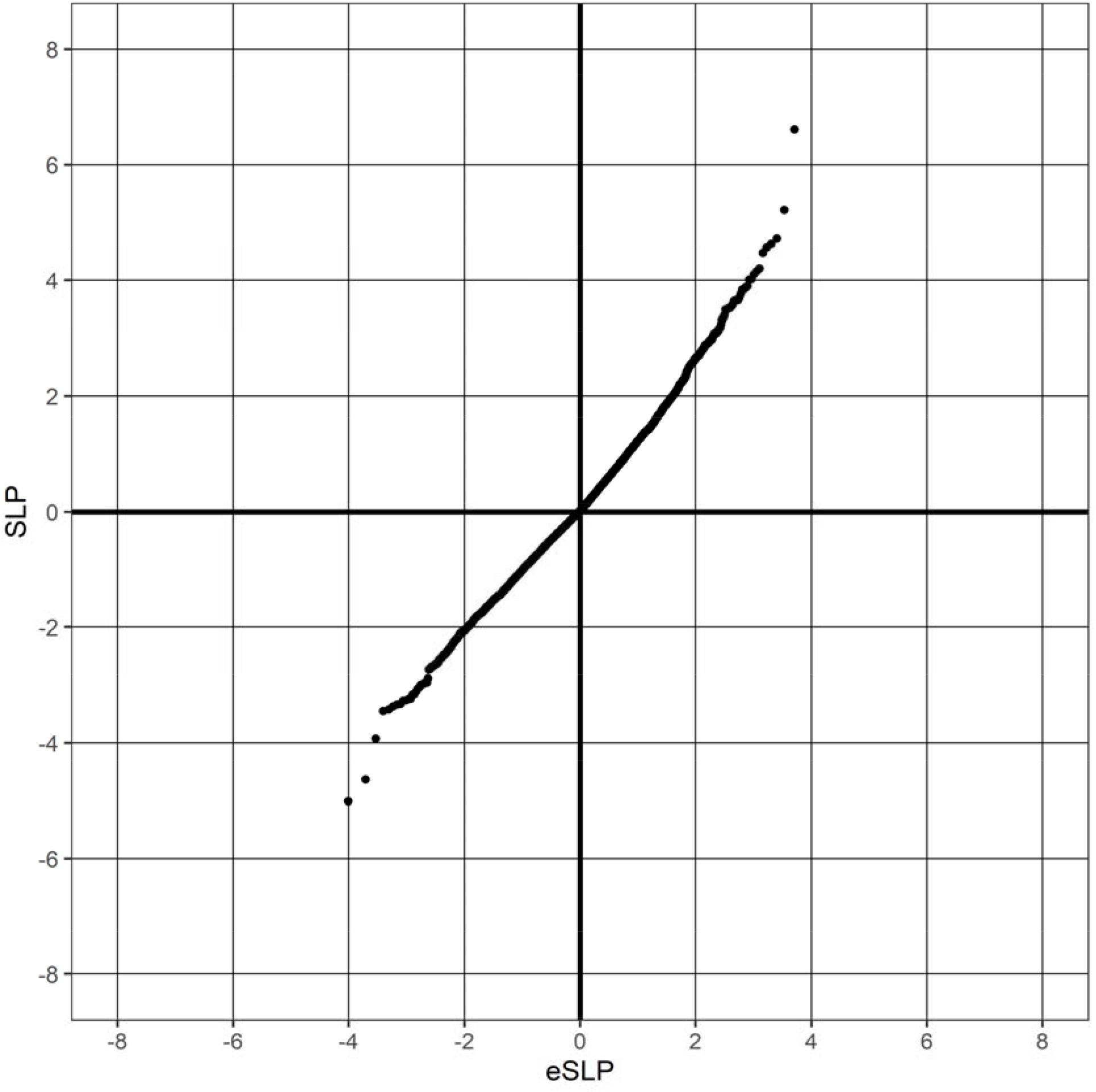
QQ plot of SLPs obtained for weighted burden analysis of association with hyperlipidaemia showing observed against expected SLP for each gene, omitting results for *MC4R*, which has SLP = 15.79.

For the two exome-wide significant genes, *MC4R* (SLP = 15.79) and *PCSK1* (SLP = 6.61), logistic regression analysis of different categories of variants was carried out to elucidate their relative contributions. The results are shown in Table 2, which shows differences between the genes relating to the implicated pattern of variants. In *MC4R*, disruptive variants (stop and frameshift) are associated with a highly significant (SLP = 6.55) increase in BMI by 2.72 units, equivalent to about 8 kg for somebody of average height, and carriers have an average BMI of 30.16. These variants occur a total of 80 times at 19 separate positions. There are no splice site variants. Additionally, nonsynonymous variants annotated by PolyPhen as probably damaging are also significantly (SLP = 4.29) associated with an average increase in BMI of 2.02 units. These occur in total 425 times at 55 positions. By contrast, other variants, including those annotated as deleterious by SIFT, are not associated with BMI changes. The estimated effect of the probably damaging variants represents an average across all the variants in this category and of course it is possible that some have major effects whereas other do not. However inspection of the detailed results showed that all of these variants were very rare (MAF<0.001) and so it was not possible to reliably assess the effect of any individual variant. In *PCSK1*, disruptive variants are also significantly (SLP=3.28) associated with an increase in BMI of 2.29 units and carriers have a mean BMI of 29.66. The estimated effect of splice site variants, which are also predicted to cause LOF, is similar, an increase of 2.01 units, but they only occur 8 times and this effect is not statistically significant. In contrast with *MC4R*, there is no suggestion that variants in *PCSK1* annotated as probably damaging have any effect on BMI. However the much larger general category of protein-altering variants is associated with a modest (0.34 units) but statistically significant (SLP = 2.74) increase in BMI. In total these occur 2,970 times, meaning that there is an average variant burden per subject of 0.015.

**Table 2A.** Results from regression analysis showing the effects on BMI of different categories of variant within the two exome-wide significant genes, *MC4R* and *PCSK1*. For each category of variant, the table shows the number of different variants of that category (at different locations) and the total number of times a variant of that category occurred. Also shown is the mean and SD of the BMI for all subjects carrying at least one variant of that category. The SLP is the signed log10 p value from the regression analysis and the estimated effect for each category is the fitted mean change in BMI after incorporating principal components and sex as covariates. Results for *MC4R*.

| Category | Number of different variants | Total number of variants | Average variant load per subject | BMI mean in carriers | BMI SD in carriers | SLP | Effect on mean BMI (95% CI) |
| --- | --- | --- | --- | --- | --- | --- | --- |
| Intronic, etc | 0 | 0 |  |  |  |  |  |
| 5 prime UTR | 25 | 286 | 0.001431 | 27.82 | 5.08 | 1.02 | 0.47 (-0.09 - 1.03) |
| Synonymous | 60 | 883 | 0.004419 | 28.14 | 5.19 | -0.66 | -0.19 (-0.50 - 0.12) |
| Splice region | 0 | 0 |  |  |  |  |  |
| 3 prime UTR | 7 | 49 | 0.000245 | 27.57 | 5.26 | 0.12 | 0.20 (-1.15 - 1.55) |
| Protein altering | 140 | 1,355 | 0.006782 | 28.24 | 5.42 | 0.03 | 0.02 (-0.34 - 0.37) |
| InDel, etc | 1 | 4 | 0.000020 | 28.76 | 8.08 | 0.33 | 1.70 (-3.03 - 6.42) |
| Disruptive | 19 | 80 | 0.000400 | 30.16 | 4.93 | 6.55 | 2.72 (1.66 - 3.79) |
| Splice site variant | 0 | 0 |  |  |  |  |  |
| Deleterious | 70 | 452 | 0.002262 | 28.70 | 5.88 | -0.54 | -0.50 (-1.44 - 0.44) |
| Possibly damaging | 23 | 201 | 0.001006 | 28.42 | 5.51 | 1.68 | 0.92 (0.13 - 1.72) |
| Probably damaging | 55 | 425 | 0.002127 | 28.98 | 5.86 | 4.29 | 2.02 (1.02 - 3.02) |
| Subjects with no variant |  | 197,329 | 0.987598 | 27.37 | 4.75 |  |  |

**Table 2B.**
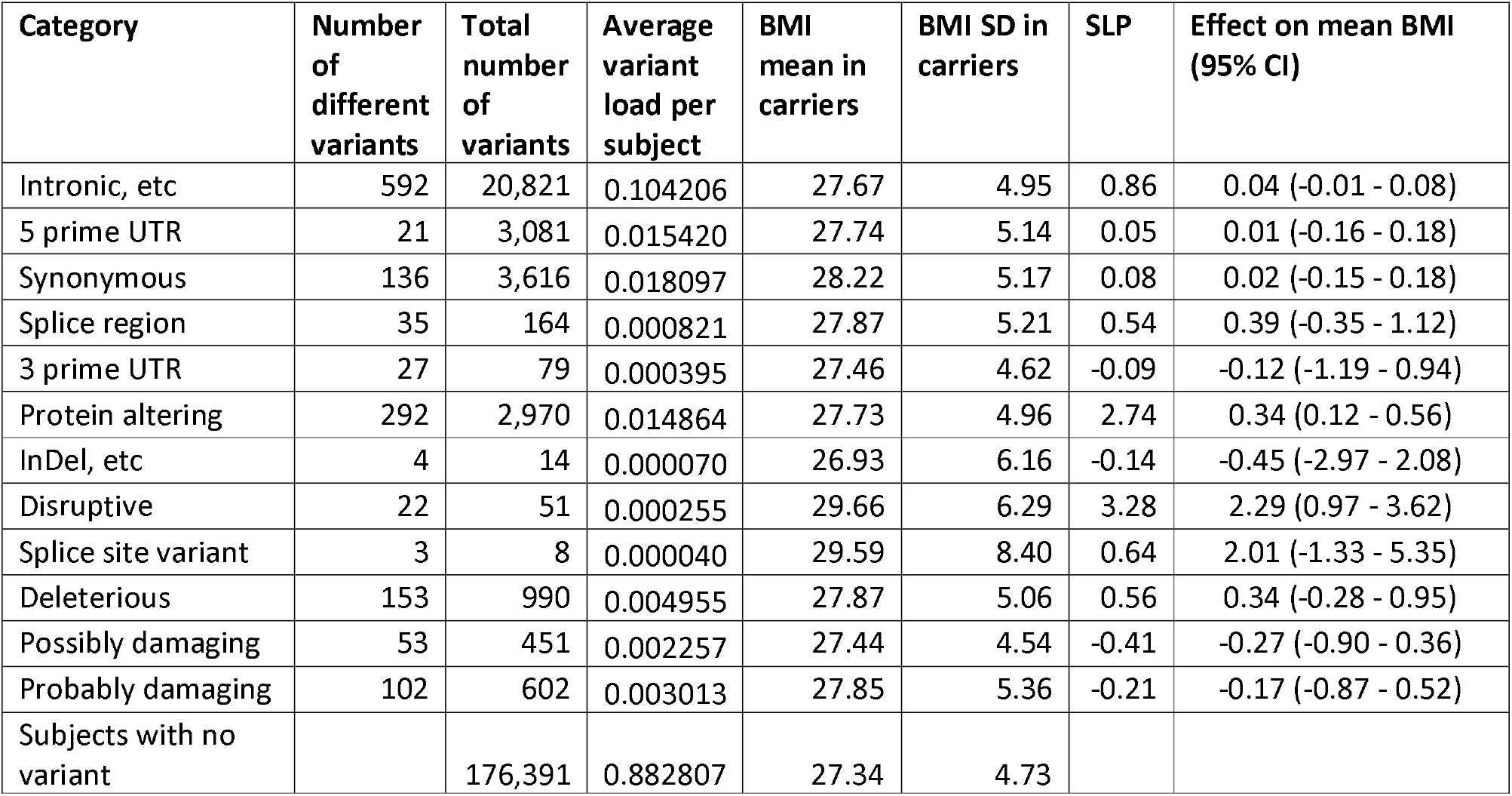
Results *for PCSK1*.

| Category | Number of different variants | Total number of variants | Average variant load per subject | BMI mean in carriers | BMI SD in carriers | SLP | Effect on mean BMI (95% CI) |
| --- | --- | --- | --- | --- | --- | --- | --- |
| Intronic, etc | 592 | 20,821 | 0.104206 | 27.67 | 4.95 | 0.86 | 0.04 (-0.01 - 0.08) |
| 5 prime UTR | 21 | 3,081 | 0.015420 | 27.74 | 5.14 | 0.05 | 0.01 (-0.16 - 0.18) |
| Synonymous | 136 | 3,616 | 0.018097 | 28.22 | 5.17 | 0.08 | 0.02 (-0.15 - 0.18) |
| Splice region | 35 | 164 | 0.000821 | 27.87 | 5.21 | 0.54 | 0.39 (-0.35 - 1.12) |
| 3 prime UTR | 27 | 79 | 0.000395 | 27.46 | 4.62 | -0.09 | -0.12 (-1.19 - 0.94) |
| Protein altering | 292 | 2,970 | 0.014864 | 27.73 | 4.96 | 2.74 | 0.34 (0.12 - 0.56) |
| InDel, etc | 4 | 14 | 0.000070 | 26.93 | 6.16 | -0.14 | -0.45 (-2.97 - 2.08) |
| Disruptive | 22 | 51 | 0.000255 | 29.66 | 6.29 | 3.28 | 2.29 (0.97 - 3.62) |
| Splice site variant | 3 | 8 | 0.000040 | 29.59 | 8.40 | 0.64 | 2.01 (-1.33 - 5.35) |
| Deleterious | 153 | 990 | 0.004955 | 27.87 | 5.06 | 0.56 | 0.34 (-0.28 - 0.95) |
| Possibly damaging | 53 | 451 | 0.002257 | 27.44 | 4.54 | -0.41 | -0.27 (-0.90 - 0.36) |
| Probably damaging | 102 | 602 | 0.003013 | 27.85 | 5.36 | -0.21 | -0.17 (-0.87 - 0.52) |
| Subjects with no variant |  | 176,391 | 0.882807 | 27.34 | 4.73 |  |  |

One would expect that by chance 20 genes would produce SLPs with absolute value greater than 3, equivalent to p < 0.001, whereas in fact there are 68, suggesting that some might have an effect on BMI while failing to reach exome-wide significance after correction for multiple testing. They are listed in Table 3. Variant category analyses were carried out for those which seemed biologically plausible as well as for genes previously reported to be causative of obesity as listed in the introduction. These analyses yielded some findings of possible interest, discussed as follows.

**Table 3.**
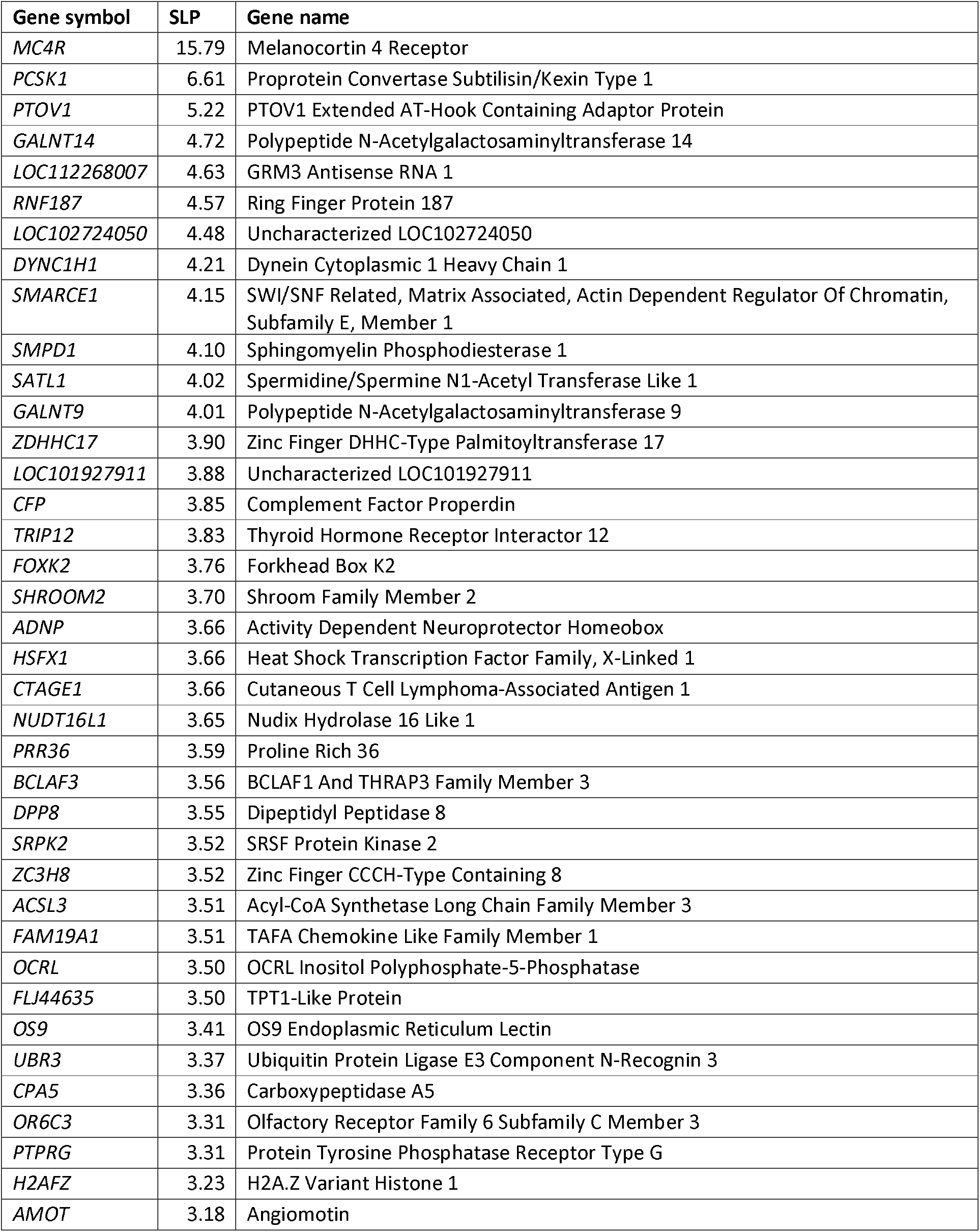
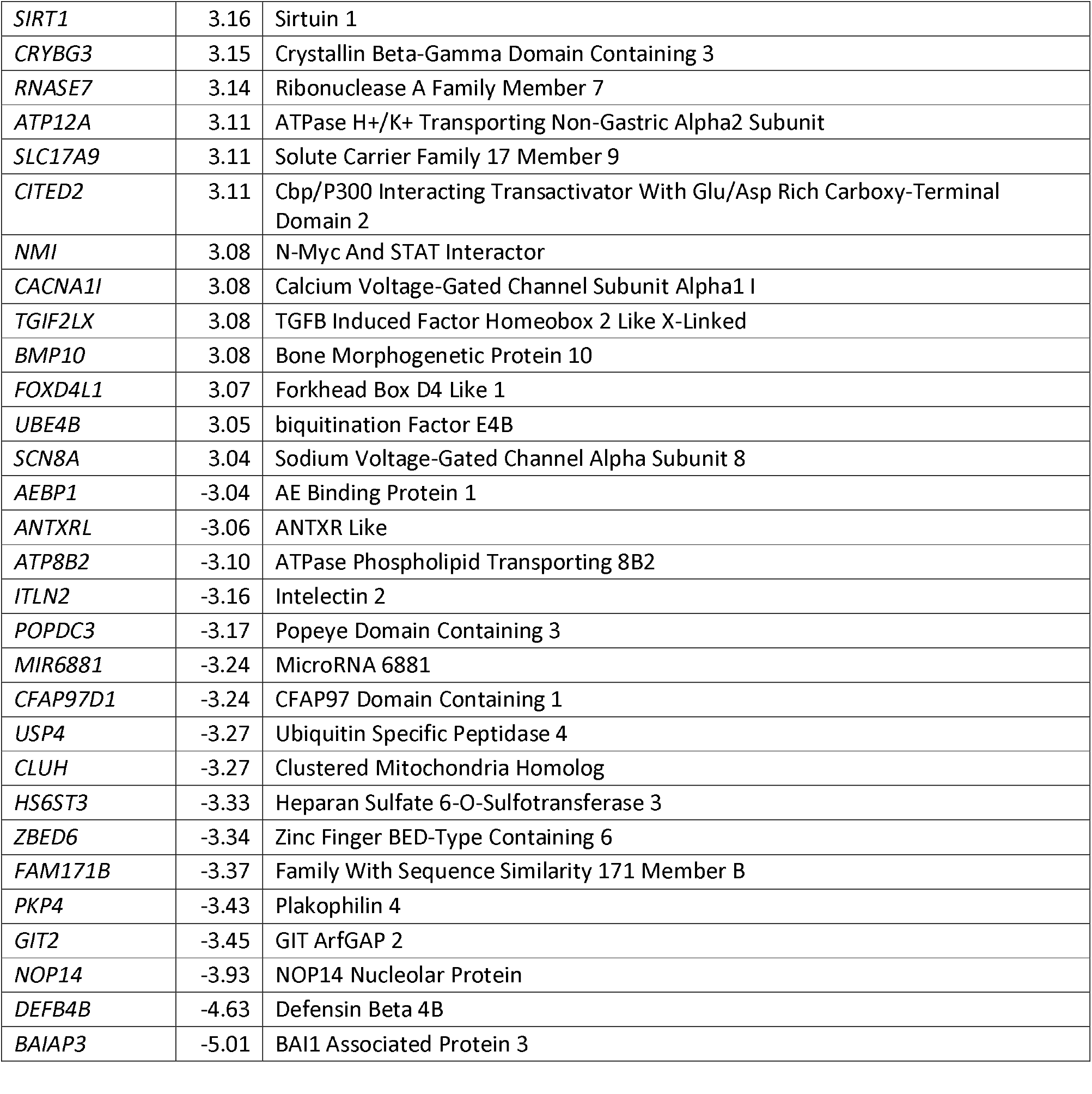
Genes with absolute value of SLP exceeding 3 or more (equivalent to p<0.001) for test of association of weighted burden score with BMI.

It is perhaps striking that two similar genes, *GALNT14* (SLP = 4.72) and *GALNT9* (SLP = 4.01), fall within the top 13 genes. These enzymes catalyze the transfer of N-acetyl-D-galactosamine (GalNAc) to the hydroxyl groups on serines and threonines in target peptides. The *GALNT9* <u>intronic SNP</u> rs11247009-A has been reported to be associated with BMI (p = 6 x 10^−9^) (Winkler et al., 2015). A study of broiler chickens claimed that in unpublished data one of the six most highly significant variants in a genome-wide study of abdominal fat was in *GALNT9* and reported that *GALNT9* expression in liver differed between lean and fat lines (Jin et al., 2017). However, overall there seems to be little prior evidence to implicate these genes as affecting BMI and they have mostly been studied in the context of cancer progression, although there is also a report of a homozygous frameshift variant of *GALNT14* being found in a patient with nonsyndromic keratoconus. The results of variant-wise analysis of these two genes are shown in Tables 4A and 4B. This shows that *GALNT14* there are 302 disruptive variants associated with a significant (SLP = 2.89) increase in BMI of 0.88 units, while in *GALNT9* there are 12 splice site variants associated with an increase in BMI of 3.97 units (SLP = 2.44) and 9 indels associated with an increase in BMI of 4.84 units (SLP = 2.65). 36 disruptive variants in *GALNT9* are also associated with an increase in BMI of 1.14 units but this is not statistically significant (SLP = 0.86).

**Table 4A.** Results from variant category regression analyses for other genes of possible interest. The tables show the numbers of variant of each category, their total numbers and the mean and SD of BMI observed in variant carriers along with the SLP and estimated effect size. Results for *GALNT14*.

| Category | Number of different variants | Total number of variants | Average variant load per subject | BMI mean in carriers | BMI SD in carriers | SLP | Effect on mean BMI (95% CI) |
| --- | --- | --- | --- | --- | --- | --- | --- |
| Intronic, etc | 877 | 20,167 | 0.100932 | 27.73 | 4.98 | -0.02 | -0.00 (-0.07 - 0.06) |
| 5 prime UTR | 38 | 697 | 0.003488 | 28.16 | 5.27 | 0.05 | 0.02 (-0.34 - 0.38) |
| Synonymous | 121 | 5558 | 0.027817 | 27.49 | 4.79 | -0.66 | -0.08 (-0.22 - 0.05) |
| Splice region | 46 | 1204 | 0.006026 | 28.94 | 5.02 | -0.33 | -0.10 (-0.38 - 0.18) |
| 3 prime UTR | 22 | 243 | 0.001216 | 27.22 | 4.71 | -0.25 | -0.17 (-0.78 - 0.43) |
| Protein altering | 299 | 9,851 | 0.049303 | 27.47 | 4.80 | 0.03 | 0.00 (-0.10 - 0.11) |
| InDel, etc | 5 | 6 | 0.000030 | 24.72 | 3.11 | -0.81 | -2.74 (-6.59 - 1.12) |
| Disruptive | 30 | 302 | 0.001511 | 28.24 | 5.35 | 2.89 | 0.88 (0.33 - 1.42) |
| Splice site variant | 13 | 50 | 0.000250 | 27.76 | 4.82 | 0.22 | 0.35 (-0.99 - 1.69) |
| Deleterious | 176 | 2,302 | 0.011521 | 27.72 | 4.91 | 1.00 | 0.32 (-0.07 - 0.71) |
| Possibly damaging | 46 | 695 | 0.003478 | 27.93 | 4.84 | 0.26 | 0.15 (-0.36 - 0.66) |
| Probably damaging | 140 | 1,335 | 0.006681 | 27.58 | 5.04 | -0.18 | -0.09 (-0.52 - 0.34) |
| Subjects with no variant |  | 171,281 | 0.857232 | 27.34 | 4.73 |  |  |

**Table 4B.** Results for *GALNT9*.

| Category | Number of different variants | Total number of variants | Average variant load per subject | BMI mean in carriers | BMI SD in carriers | SLP | Effect on mean BMI (95% CI) |
| --- | --- | --- | --- | --- | --- | --- | --- |
| Intronic, etc | 613 | 25,032 | 0.125281 | 27.69 | 4.99 | -1.08 | -0.05 (-0.10 - 0.01) |
| 5 prime UTR | 34 | 132 | 0.000661 | 28.21 | 4.57 | -0.24 | -0.22 (-1.01 - 0.57) |
| Synonymous | 183 | 10,134 | 0.050719 | 27.90 | 5.20 | 0.69 | 0.06 (-0.04 - 0.17) |
| Splice region | 44 | 231 | 0.001156 | 27.45 | 4.87 | 0.24 | 0.17 (-0.45 - 0.79) |
| 3 prime UTR | 228 | 11,273 | 0.056419 | 27.82 | 5.00 | -0.12 | -0.01 (-0.10 - 0.08) |
| Protein altering | 362 | 2,952 | 0.014774 | 27.66 | 4.95 | 0.43 | 0.13 (-0.16 - 0.41) |
| InDel, etc | 5 | 9 | 0.000045 | 32.21 | 10.83 | 2.65 | 4.84 (1.68 - 8.01) |
| Disruptive | 19 | 36 | 0.000180 | 28.17 | 7.12 | 0.86 | 1.14 (-0.40 - 2.68) |
| Splice site variant | 7 | 12 | 0.000060 | 31.20 | 12.28 | 2.44 | 3.97 (1.24 - 6.70) |
| Deleterious | 207 | 1,626 | 0.008138 | 27.63 | 4.99 | -0.08 | -0.04 (-0.45 - 0.37) |
| Possibly damaging | 83 | 951 | 0.004760 | 27.68 | 4.79 | 0.46 | 0.21 (-0.24 - 0.66) |
| Probably damaging | 109 | 736 | 0.003684 | 27.69 | 5.05 | 0.50 | 0.25 (-0.25 - 0.75) |
| Subjects with no variant |  | 172,467 | 0.863168 | 27.34 | 4.73 |  |  |

The results for *SIRT1* (SLP = 3.16) are potentially of interest because SIRT1 and other sirtuins have effects similar to calorie restriction and reduced expression of *SIRT1* and *SIRT2* promotes adipogenesis and accumulation of visceral fat (Chang and Guarente, 2014; Perrini et al., 2020). From these findings one might well predict that genetic variants damaging *SIRT1* might lead to increased BMI. The results from variant-wise analysis are shown in Table 4C, which shows only weakly significant effects from disruptive (SLP = 1.34) and possibly damaging (SLP = 1.61) variants.

**Table 4C.**
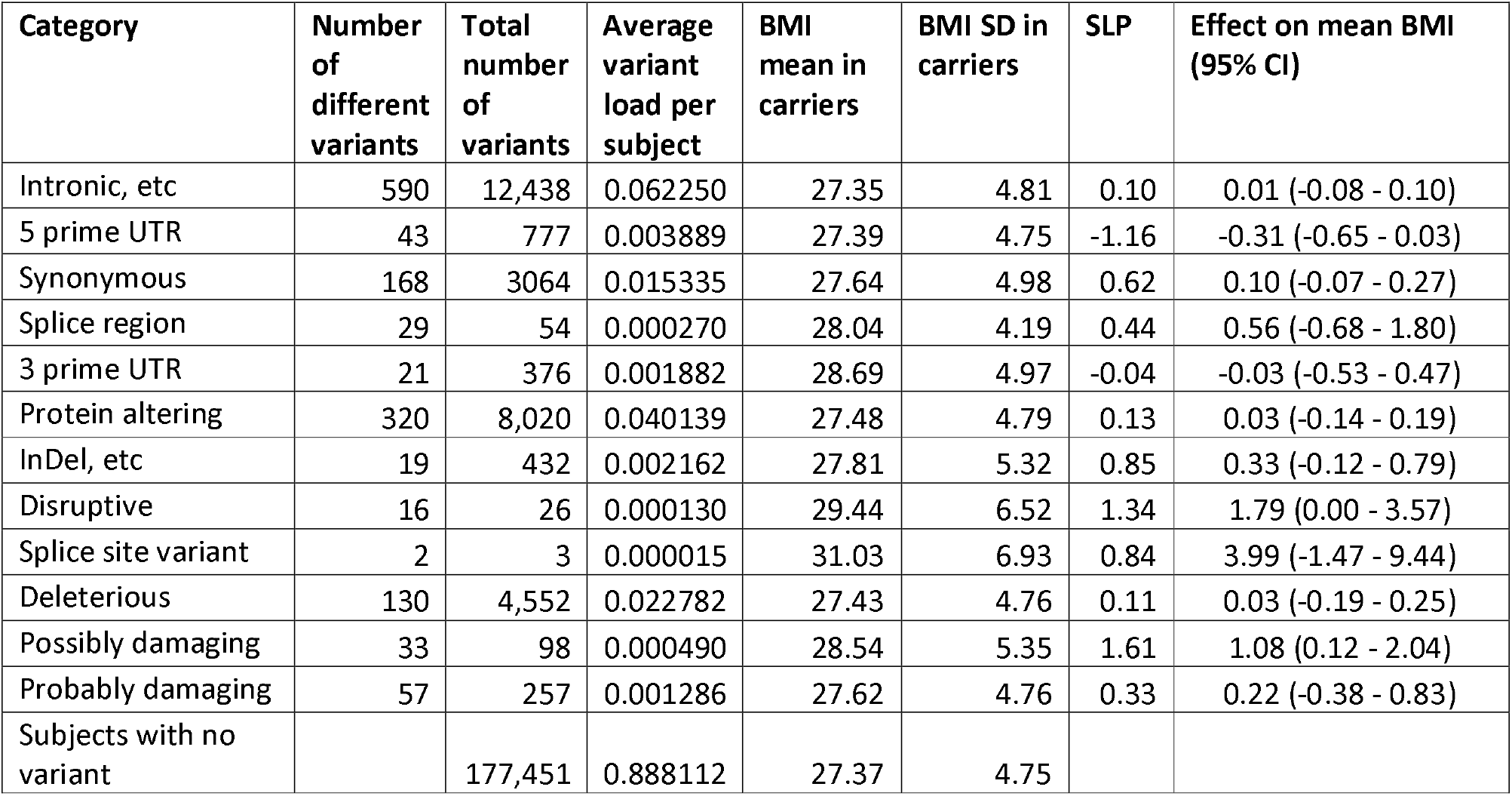
Results for *SIRT1*.

| Category | Number of different variants | Total number of variants | Average variant load per subject | BMI mean in carriers | BMI SD in carriers | SLP | Effect on mean BMI (95% CI) |
| --- | --- | --- | --- | --- | --- | --- | --- |
| Intronic, etc | 590 | 12,438 | 0.062250 | 27.35 | 4.81 | 0.10 | 0.01 (-0.08 - 0.10) |
| 5 prime UTR | 43 | 777 | 0.003889 | 27.39 | 4.75 | -1.16 | -0.31 (-0.65 - 0.03) |
| Synonymous | 168 | 3064 | 0.015335 | 27.64 | 4.98 | 0.62 | 0.10 (-0.07 - 0.27) |
| Splice region | 29 | 54 | 0.000270 | 28.04 | 4.19 | 0.44 | 0.56 (-0.68 - 1.80) |
| 3 prime UTR | 21 | 376 | 0.001882 | 28.69 | 4.97 | -0.04 | -0.03 (-0.53 - 0.47) |
| Protein altering | 320 | 8,020 | 0.040139 | 27.48 | 4.79 | 0.13 | 0.03 (-0.14 - 0.19) |
| InDel, etc | 19 | 432 | 0.002162 | 27.81 | 5.32 | 0.85 | 0.33 (-0.12 - 0.79) |
| Disruptive | 16 | 26 | 0.000130 | 29.44 | 6.52 | 1.34 | 1.79 (0.00 - 3.57) |
| Splice site variant | 2 | 3 | 0.000015 | 31.03 | 6.93 | 0.84 | 3.99 (-1.47 - 9.44) |
| Deleterious | 130 | 4,552 | 0.022782 | 27.43 | 4.76 | 0.11 | 0.03 (-0.19 - 0.25) |
| Possibly damaging | 33 | 98 | 0.000490 | 28.54 | 5.35 | 1.61 | 1.08 (0.12 - 2.04) |
| Probably damaging | 57 | 257 | 0.001286 | 27.62 | 4.76 | 0.33 | 0.22 (-0.38 - 0.83) |
| Subjects with no variant |  | 177,451 | 0.888112 | 27.37 | 4.75 |  |  |

*ZBED6* (SLP = −3.33) codes for a transcriptional inhibitor of *IGF2* which has a major impact on muscle development in placental mammals and CRISPR/Cas9 disruption of its binding site is being used commercially to produce strains of pigs which are leaner and have enhanced muscle development (Liu et al., 2019; Younis et al., 2018). The results for variant-wise analysis are shown in Table 4D, showing that disruptive variants are associated with a reduction in BMI of 1.59 units (SLP = −2.48) and deleterious nonsynonymous variants with a reduction of 0.37 units (SLP = −1.49).

The gene with the most negative SLP, *BAIAP3* (SLP = −5.01), may have some role in insulin secretion but does not in general seem to be an obvious candidate to have effects on BMI (Zhang et al., 2017). Splice site variants are associated with a reduction in BMI of 1.41 units (SLP = −3.47).

**Table 4D.** Results for *ZBED6*.

| Category | Number of different variants | Total number of variants | Average variant load per subject | BMI mean in carriers | BMI SD in carriers | SLP | Effect on mean BMI (95% CI) |
| --- | --- | --- | --- | --- | --- | --- | --- |
| Intronic, etc | 0 | 0 |  |  |  |  |  |
| 5 prime UTR | 0 | 0 |  |  |  |  |  |
| Synonymous | 145 | 1,316 | 0.006586 | 28.12 | 5.36 | 0.39 | 0.11 (-0.15 - 0.37) |
| Splice region | 0 | 0 |  |  |  |  |  |
| 3 prime UTR | 19 | 104 | 0.000521 | 28.26 | 4.34 | 1.24 | 0.88 (-0.05 - 1.81) |
| Protein altering | 322 | 4,785 | 0.023948 | 27.44 | 4.80 | -0.07 | -0.02 (-0.18 - 0.15) |
| InDel, etc | 11 | 102 | 0.000510 | 27.13 | 4.54 | -0.17 | -0.20 (-1.13 - 0.74) |
| Disruptive | 40 | 74 | 0.000370 | 25.72 | 3.48 | -2.48 | -1.59 (-2.68 - -0.51) |
| Splice site variant | 0 | 0 |  |  |  |  |  |
| Deleterious | 121 | 914 | 0.004574 | 27.36 | 4.92 | -1.49 | -0.37 (-0.72 - -0.02) |
| Possibly damaging | 69 | 410 | 0.002052 | 27.68 | 4.57 | 0.25 | 0.14 (-0.35 - 0.63) |
| Probably damaging | 99 | 451 | 0.002257 | 27.44 | 4.61 | -0.01 | -0.01 (-0.48 - 0.47) |
| Subjects with no variant |  | 193,605 | 0.968960 | 27.37 | 4.75 |  |  |

**Table 4E.** Results for *BDNF*.

| Category | Number of different variants | Total number of variants | Average variant load per subject | BMI mean in carriers | BMI SD in carriers | SLP | Effect on mean BMI (95% CI) |
| --- | --- | --- | --- | --- | --- | --- | --- |
| Intronic, etc | 1,002 | 40,883 | 0.204612 | 27.56 | 4.90 | 0.36 | 0.02 (-0.03 - 0.06) |
| 5 prime UTR | 346 | 14,309 | 0.071614 | 27.77 | 5.05 | 1.33 | 0.08 (-0.00 - 0.17) |
| Synonymous | 55 | 1,150 | 0.005756 | 27.58 | 5.09 | 0.15 | 0.05 (-0.23 - 0.33) |
| Splice region | 23 | 2,234 | 0.011181 | 27.21 | 4.72 | -0.82 | -0.14 (-0.34 - 0.06) |
| 3 prime UTR | 325 | 6,410 | 0.032081 | 27.64 | 5.02 | 0.25 | 0.03 (-0.08 - 0.15) |
| Protein altering | 97 | 1,910 | 0.009559 | 27.53 | 4.78 | 0.56 | 0.23 (-0.19 - 0.64) |
| InDel, etc | 1 | 2 | 0.000010 | 26.16 | 5.03 | -0.10 | -0.83 (-7.52 - 5.85) |
| Disruptive | 6 | 325 | 0.001627 | 27.53 | 4.74 | 0.28 | 0.17 (-0.36 - 0.69) |
| Splice site variant | 9 | 33 | 0.000165 | 26.61 | 4.85 | -0.37 | -0.65 (-2.29 - 1.00) |
| Deleterious | 54 | 296 | 0.001481 | 27.53 | 4.92 | -0.01 | -0.01 (-0.60 - 0.59) |
| Possibly damaging | 13 | 64 | 0.000320 | 28.27 | 5.56 | 0.56 | 0.68 (-0.56 - 1.93) |
| Probably damaging | 31 | 1,279 | 0.006401 | 27.50 | 4.77 | -0.16 | -0.09 (-0.57 - 0.39) |
| Subjects with no variant |  | 151,624 | 0.758852 | 27.33 | 4.72 |  |  |

**Table 4F.** Results for *NPC2*.

| Category | Number of different variants | Total number of variants | Average variant load per subject | BMI mean in carriers | BMI SD in carriers | SLP | Effect on mean BMI (95% CI) |
| --- | --- | --- | --- | --- | --- | --- | --- |
| Intronic, etc | 118 | 2,755 | 0.013788 | 27.50 | 4.81 | 0.41 | 0.08 (-0.10 - 0.26) |
| 5 prime UTR | 41 | 361 | 0.001807 | 27.75 | 4.57 | -1.54 | -0.55 (-1.05 - -0.05) |
| Synonymous | 35 | 299 | 0.001496 | 26.68 | 4.53 | -1.51 | -0.59 (-1.14 - -0.04) |
| Splice region | 11 | 907 | 0.004539 | 27.79 | 4.98 | -0.10 | -0.04 (-0.36 - 0.28) |
| 3 prime UTR | 85 | 1,461 | 0.007312 | 27.91 | 4.89 | 0.46 | 0.12 (-0.13 - 0.37) |
| Protein altering | 72 | 1,573 | 0.007873 | 27.69 | 4.96 | -0.43 | -0.18 (-0.58 - 0.22) |
| InDel, etc | 1 | 1 | 0.000005 | 23.91 |  | -0.28 |  |
| Disruptive | 10 | 111 | 0.000556 | 28.94 | 5.74 | 1.27 | 0.87 (-0.03 - 1.76) |
| Splice site variant | 3 | 3,119 | 0.015610 | 27.63 | 4.95 | 3.10 | 0.28 (0.11 - 0.45) |
| Deleterious | 41 | 742 | 0.003714 | 28.45 | 5.10 | 0.96 | 0.40 (-0.10 - 0.90) |
| Possibly damaging | 12 | 525 | 0.002628 | 26.97 | 4.56 | -0.68 | -0.33 (-0.86 - 0.19) |
| Probably damaging | 19 | 68 | 0.000340 | 27.51 | 4.59 | -0.08 | -0.13 (-1.33 - 1.07) |
| Subjects with no variant |  | 189,490 | 0.948365 | 27.36 | 4.75 |  |  |

It is well established that variants in *LEP* (SLP = 0.61) and *LEPR* (SLP = 0.13) can cause obesity but the gene-based analyses produced no evidence to implicate them. The results of variant-wise analyses are shown in Tables 5A and 5B. It can be seen that subjects with disruptive and splice site variants in *LEP* do indeed have substantially higher BMIs but because there are only 6 of them this does not produce a statistically significant effect, at least if one corrects for the numbers of categories tested. There is no suggestion that any other type of variant has an effect. By contrast, in *LEPR* there are a total of 88 disruptive and splice site variants but their effect on mean BMI is negligible, as is also the case for other types of variant.

**Table 5A.** Results from variant category regression analyses for *LEP* and *LEPR*. The tables show the numbers of variant of each category, their total numbers and the mean and SD of BMI observed in variant carriers along with the SLP and estimated effect size. Results for *LEP*.

| Category | Number of different variants | Total number of variants | Average variant load per subject | BMI mean in carriers | BMI SD in carriers | SLP | Effect on mean BMI (95% CI) |
| --- | --- | --- | --- | --- | --- | --- | --- |
| Intronic, etc | 58 | 7,313 | 0.036600 | 27.61 | 5.00 | -0.16 | -0.02 (-0.13 - 0.09) |
| 5 prime UTR | 4 | 13 | 0.000065 | 29.44 | 5.61 | 1.00 | 2.16 (-0.46 - 4.78) |
| Synonymous | 45 | 1,153 | 0.005771 | 27.77 | 5.13 | 1.71 | 0.32 (0.05 - 0.60) |
| Splice region | 2 | 4 | 0.000020 | 30.91 | 6.67 | 0.83 | 3.41 (-1.31 - 8.14) |
| 3 prime UTR | 10 | 2,486 | 0.012442 | 27.63 | 4.85 | -0.20 | -0.05 (-0.24 - 0.14) |
| Protein altering | 50 | 1,235 | 0.006181 | 28.62 | 5.34 | -0.21 | -0.07 (-0.37 - 0.22) |
| InDel, etc | 1 | 1 | 0.000005 | 27.70 |  | -0.01 | -0.14 (-9.59 - 9.30) |
| Disruptive | 4 | 5 | 0.000025 | 32.05 | 3.90 | 1.64 | 4.80 (0.58 - 9.03) |
| Splice site variant | 1 | 1 | 0.000005 | 33.46 |  | 0.68 | 5.91 (-3.54 - 15.35) |
| Deleterious | 18 | 59 | 0.000295 | 27.10 | 4.00 | -0.48 | -0.96 (-2.95 - 1.03) |
| Possibly damaging | 8 | 91 | 0.000455 | 27.30 | 4.91 | 0.01 | 0.01 (-1.06 - 1.09) |
| Probably damaging | 15 | 49 | 0.000245 | 27.27 | 4.05 | 0.42 | 0.95 (-1.21 - 3.11) |
| Subjects with no variant |  | 188,116 | 0.941489 | 27.36 | 4.74 |  |  |

**Table 5B.** Results for *LEPR*.

| Category | Number of different variants | Total number of variants | Average variant load per subject | BMI mean in carriers | BMI SD in carriers | SLP | Effect on mean BMI (95% CI) |
| --- | --- | --- | --- | --- | --- | --- | --- |
| Intronic, etc | 2,481 | 51,218 | 0.256337 | 27.43 | 4.80 | -0.65 | -0.02 (-0.06 - 0.02) |
| 5 prime UTR | 48 | 3,200 | 0.016015 | 27.25 | 4.73 | -0.01 | -0.00 (-0.17 - 0.17) |
| Synonymous | 145 | 4,298 | 0.021511 | 27.67 | 4.96 | -0.13 | -0.02 (-0.18 - 0.13) |
| Splice region | 43 | 147 | 0.000736 | 27.11 | 4.57 | -0.22 | -0.20 (-0.98 - 0.58) |
| 3 prime UTR | 19 | 117 | 0.000586 | 28.26 | 5.55 | 1.24 | 0.83 (-0.05 - 1.70) |
| Protein altering | 396 | 3,862 | 0.019329 | 27.64 | 4.82 | -0.26 | -0.07 (-0.32 - 0.17) |
| InDel, etc | 3 | 4 | 0.000020 | 27.31 | 4.59 | 0.02 | 0.14 (-4.58 - 4.86) |
| Disruptive | 25 | 53 | 0.000265 | 27.49 | 5.13 | 0.06 | 0.11 (-1.19 - 1.40) |
| Splice site variant | 8 | 35 | 0.000175 | 26.47 | 5.69 | -0.47 | -0.77 (-2.37 - 0.83) |
| Deleterious | 159 | 1,950 | 0.009759 | 27.76 | 4.96 | 0.11 | 0.05 (-0.30 - 0.40) |
| Possibly damaging | 77 | 990 | 0.004955 | 27.55 | 4.77 | 0.57 | 0.21 (-0.17 - 0.60) |
| Probably damaging | 83 | 1,189 | 0.005951 | 27.96 | 4.99 | 0.31 | 0.14 (-0.27 - 0.55) |
| Subjects with no variant |  | 153,180 | 0.766640 | 27.36 | 4.74 |  |  |

A common nonsynonymous variant *BDNF*, rs6265, causes a Val66Met substitution which was originally reported to be associated with anorexia nervosa and minimum BMI in anorexia nervosa patients and whose effect on BMI was subsequently confirmed in large GWAS samples (Pulit et al., 2019; Ribasés et al., 2003). This variant shows highly significant association in the current sample (SLP = −21.86). The number of subjects with Val/Val, Val/Met and Met/Met genotypes is 132,003, 60,639 and 7,165 with uncorrected mean BMIs of 27.47, 27.22 and 26.96. The per-allele effect size on BMI as estimated from multiple linear regression analysis including principal components and sex as covariates is −0.19 (−0.23 – −0.15). However the gene-wise weighted burden analysis of *BDNF* using rare variants produced no evidence for association (SLP = 0.41) and variant-wise analyses likewise failed to show any effect from any category of rare variant. The mean effect size for protein-altering variants was 0.23 but there were only 1,910 of these in total and the result does not approach statistical significance. These results are shown in Table 4E.

Other genes previously implicated in obesity which likewise failed to show evidence of association in either gene-wise analyses or variant category analyses include *SIM1* (SLP = 0.89), *NTRK2* (SLP = 0.88), *KSR2* (SLP = 0.17), *CPE* (SLP = −0.35), *SH2B1* (SLP = 0.78), *TUB* (SLP = −0.08) and *FTO* (SLP = 1.02).

In order to see if any additional genes were highlighted by analysing gene sets, gene set analysis was performed as described above after first removing all genes with absolute SLP value greater than 3. In order to correct for the observed inflation of the positive SLPs, the absolute value of each SLP was divided by an average inflation factor of 1.13 before being utilised to contribute to the set-wise chi-squared statistic. Following this adjustment, no gene set produced a result significant after correction for multiple testing. The highest MLP was 2.45, achieved by the set SPECIFIC TRANSCRIPTIONAL REPRESSOR ACTIVITY. Out of 1,454 sets, the fifth highest ranked was REGULATION OF LIPID METABOLIC PROCESS (MLP = 1.93). This contains 12 genes including *NPC2* (SLP = 2.80), which is involved in cholesterol transport and recessively acting variants in *NPC2* are a cause of Niemann-Pick C disease in which lipid accumulation causes neurodegeneration (Xu et al., 2019). NPC2 presents cholesterol to NPC1 and rare LOF variants in *NPC1* are known to cause obesity although *NPC1* does not demonstrate association with BMI in the current sample (SLP = 0.15) (Liu et al., 2017). In a GWAS of obesity in F2 pigs a variant within *NPC2*, rs81396056, produced the most highly significant result (p = 9 x 10^−17^) (Kogelman et al., 2014). The results of variant category analysis of *NPC2* are shown in Table 4F and it can be seen that there is significant (SLP = 3.10) association of 3,119 splice site variants, occurring at 3 different positions, with an average increase in BMI of 0.28. Disruptive variants are also associated with higher BMI but there are only 111 of them and this result is not statistically significant.

Results for all genes and gene sets along with variant category analyses for selected genes are presented in the Supplementary Tables S1, S2 and S3.

## Discussion

These analyses help to elucidate the impact of rare genetic variants on a complex phenotype such as BMI and also illustrate some of the challenges of dealing with exome sequence data. The gene-wise weighted burden analyses successfully identify two genes already known to impact BMI, *MC4R* and *PCSK1*, but fail to detect effects of other known obesity genes. In due course sequence data will become available for all 500,000 UK Biobank participants and it is reasonable to expect that this larger dataset will produce additional results. For example, the subjects with LOF variants in *LEP* do have notably higher BMIs but there are so few of them that they do not produce a statistically significant result in this sample. Obviously, the power to detect association depends both on the effect size and the frequency of variants, and power will improve with increased sample size. To take another example of this issue, although the results for the *BDNF* Val66Met variant are highly statistically significant, other protein altering variants in *BDNF* are associated with a larger average effect size but do not produce a statistically significant result because they are cumulatively so much rarer.

The results provide some indication about the quantitative effects of sequence variants but we should first note that the UK Biobank is not completely representative. It consists of volunteer participants who are on average older and healthier than the population as a whole. One implication of this is that subjects with more severe phenotypes will be less likely to be included and an overall effect of this will be to underestimate the effect size of rare variants which can cause morbidity and premature mortality. For example, we can observe that LOF variants in *MC4R* and *PCSK1* are associated with an average increase of 2 or more BMI units but that this estimate may well represent a floor for the real effect size.

The public health impact of genetic variants depends on their effect and on how many people carry them. For those categories of variant which are rare, the proportion of people carrying such a variant will be approximated by the average variant load because few people will have more than one variant. Thus, we may say that 0.04% of this sample has a LOF variant in *MC4R* associated with an increase of 2.7 in BMI while 0.2% have a variant annotated as probably damaging by PolyPhen associated with an average BMI increase of 2.0. Likewise, less than 0.03% of the sample has a LOF variant in *PCSK1* which tends to increase BMI by 2.3 units whereas 1.5% carry a protein altering variant associated with an average BMI increase of 0.3.

The analyses fail to conclusively implicate novel genes as influencing BMI. The three which are arguably biologically the most plausible are *SIRT1, ZBED6* and *NPC2* but it must be acknowledged that the statistical evidence supporting their involvement is fairly weak. Conversely, there are other genes with higher statistical significance but whose function, as far as it is known, does not immediately suggest that they would have a prominent role in influencing BMI. It is clear that additional data will be needed to arrive at definitive solutions, whether it be from the remaining UK Biobank subjects or from alternative sources.

The results from these analyses would seem to point to very rare variants in a fairly small number of genes as having detectable effects on BMI but there are some caveats which are worth stating. Firstly, the approach used is based on the assumption that when variants are considered jointly then they will tend to have the same direction of effect on the phenotype. This assumption might seem to be reasonable for LOF variants, expected to reduce the functioning of a gene, but the method would fail if some non-synonymous variants reduced function but were balanced out by others which produced gain of function. While we may expect that on average a non-synonymous change, especially one annotated as damaging or deleterious, will be more likely to impair than improve function it is important to acknowledge that if there is a good deal of heterogeneity of effect then genes and classes of variant will fail to achieve statistical significance. Given this consideration, these results should not be taken to exclude the possibility that there may be very large numbers of individually rare variants in many genes which might cumulatively make a substantial contribution to the overall variance of BMI in the population.

Another point to make is that association studies such as this, especially those based on population samples, are not expected to necessarily identify genes which most strongly influence BMI but rather genes in which there is substantial naturally occurring variation which affects BMI. For example, there are large variations in the frequency with which LOF variants are observed in different genes, reflecting partly the size of the gene but also selection pressures. Only 6 subjects have LOF variants in *LEP* compared to thousands in *NPC2* and so *LEP* does not produce a detectable signal. However it may well be that the recognition of *LEP* as potentially having a major and direct impact on BMI would lead on to functional studies which could yield useful understanding of the underlying physiology. It should be noted that the selection pressures reducing variation in a particular gene might relate to the phenotype under consideration, here BMI, but might also involve other biological processes impacting on fitness.

The current study is intended to provide an overview of the relationship between rare variants and BMI and there is plenty of scope for more detailed analyses. These might include investigating the relationship between rare variants and GWAS signals, investigation of particular variants on predicted protein structure and function and relationships between genetic and environmental effects.

To conclude, the study of very large, exome-sequenced samples such as the UK Biobank can afford us further insights into the relationship between genetic variation and a quantitative, health-related phenotype such as BMI. However even such large samples may fail to produce definitive results and there is a good case to be made for increasing the available data still further.

## Data Availability

The raw data is available on application to UK Biobank. Detailed results with variant counts cannot be made available because they might be used for subject identification. Scripts and relevant derived variables will be deposited in UK Biobank. Software and scripts used to carry out the analyses are available at https://github.com/davenomiddlenamecurtis.

## Acknowledgments

This research has been conducted using the UK Biobank Resource. The author wishes to acknowledge the staff supporting the High Performance Computing Cluster, Computer Science Department, University College London. This work was carried out in part using resources provided by BBSRC equipment grant BB/R01356X/1. The author wishes to thank the participants who volunteered for the UK Biobank project.

## References

Adzhubei, I., Jordan, D.M., Sunyaev, S.R. (2013) Predicting functional effect of human missense mutations using PolyPhen-2. Curr. Protoc. Hum. Genet. 7 Unit7.20.

Chang, C.C., Chow, C.C., Tellier, L.C., Vattikuti, S., Purcell, S.M., Lee, J.J. (2015) Second-generation PLINK: rising to the challenge of larger and richer datasets. Gigascience 4, 7.

Chang, H.C., Guarente, L. (2014) SIRT1 and other sirtuins in metabolism. Trends Endocrinol. Metab.

Curtis, D. (2012) A rapid method for combined analysis of common and rare variants at the level of a region, gene, or pathway. Adv Appl Bioinform Chem 5, 1–9.

Curtis, D. (2016) Pathway analysis of whole exome sequence data provides further support for the involvement of histone modification in the aetiology of schizophrenia. Psychiatr. Genet. 26, 223–7.

Curtis, D. (2021a) Multiple Linear Regression Allows Weighted Burden Analysis of Rare Coding Variants in an Ethnically Heterogeneous Population. Hum. Hered. 1–10.

Curtis, D. (2021b) Analysis of 200,000 exome-sequenced UK Biobank subjects illustrates the contribution of rare genetic variants to hyperlipidaemia. medRxiv.

Curtis, D. (2021c) Weighted burden analysis in 200,000 exome-sequenced subjects characterises rare variant effects on risk of type 2 diabetes. medRxiv 2021.01.08.21249453.

Galinsky, K.J., Bhatia, G., Loh, P.R., Georgiev, S., Mukherjee, S., Patterson, N.J., Price, A.L. (2016) Fast Principal-Component Analysis Reveals Convergent Evolution of ADH1B in Europe and East Asia. Am. J. Hum. Genet. 98, 456–472.

Jin, P., Wu, X., Xu, S., Zhang, H., Li, Y., Cao, Z., Li, H., Wang, S. (2017) Differential expression of six genes and correlation with fatness traits in a unique broiler population. Saudi J. Biol. Sci. 24, 945–949.

Kogelman, L.J.A., Pant, S.D., Fredholm, M., Kadarmideen, H.N. (2014) Systems genetics of obesity in an F2 pig model by genome-wide association, genetic network, and pathway analyses. Front. Genet. 5.

Kumar, P., Henikoff, S., Ng, P.C. (2009) Predicting the effects of coding non-synonymous variants on protein function using the SIFT algorithm. Nat. Protoc. 4, 1073–1081.

Liu, R., Zou, Y., Hong, J., Cao, M., Cui, B., Zhang, H., Chen, M., Shi, J., Ning, T., Zhao, S., Liu, W., Xiong, H., Wei, C., Qiu, Z., Gu, W., Zhang, Y., Li, W., Miao, L., Sun, Y., Yang, M., Wang, R., Ma, Q., Xu, M., Xu, Y., Wang, T., Katie Chan, K.H., Zuo, X., Chen, H., Qi, L., Lai, S., Duan, S., Song, B., Bi, Y., Liu, S., Wang, W., Ning, G., Wang, J. (2017) Rare loss-of-function variants in npc1 predispose to human obesity. Diabetes 66, 935–947.

Liu, Xiaofeng, Liu, H., Wang, M., Li, R., Zeng, J., Mo, D., Cong, P., Liu, Xiaohong, Chen, Y., He, Z. (2019) Disruption of the ZBED6 binding site in intron 3 of IGF2 by CRISPR/Cas9 leads to enhanced muscle development in Liang Guang Small Spotted pigs. Transgenic Res. 28, 141–150.

McLaren, W., Gil, L., Hunt, S.E., Riat, H.S., Ritchie, G.R.S., Thormann, A., Flicek, P., Cunningham, F. (2016) The Ensembl Variant Effect Predictor. Genome Biol. 17, 122.

Müller, M.J., Geisler, C., Blundell, J., Dulloo, A., Schutz, Y., Krawczak, M., Bosy-Westphal, A., Enderle, J., Heymsfield, S.B. (2018) The case of GWAS of obesity: does body weight control play by the rules? Int. J. Obes.

Perrini, S., Porro, S., Nigro, Pasquale, Cignarelli, A., Caccioppoli, C., Valentina Genchi, A., Martines, G., De Fazio, M., Capuano, P., Natalicchio, A., Laviola, Luigi, Giorgino, F. (2020) Reduced SIRT1 and SIRT2 expression promotes adipogenesis of human visceral adipose stem cells and associates with accumulation of visceral fat in human obesity. Int. J. Obes. 44, 307–319.

Pulit, S.L., Stoneman, C., Morris, A.P., Wood, A.R., Glastonbury, C.A., Tyrrell, J., Yengo, L., Ferreira, T., Marouli, E., Ji, Y., Yang, J., Jones, S., Beaumont, R., Croteau-Chonka, D.C., Winkler, T.W., Hattersley, A.T., Loos, R.J.F., Hirschhorn, J.N., Visscher, P.M., Frayling, T.M., Yaghootkar, H., Lindgren, C.M. (2019) Meta-Analysis of genome-wide association studies for body fat distribution in 694 649 individuals of European ancestry. Hum. Mol. Genet. 28, 166–174.

R Core Team (2014) R: A language and environment for statistical computing. R Foundation for Statistical Computing, Vienna, Austria., Austria.

Ribasés, M., Gratacòs, M., Armengol, L., De Cid, R., Badía, A., Jiménez, L., Solano, R., Vallejo, J., Fernández, F., Estivill, X. (2003) Met66 in the brain-derived neurotrophic factor (BDNF) precursor is associated with anorexia nervosa restrictive type. Mol. Psychiatry 8, 745–751.

Stutzmann, F., Vatin, V., Cauchi, S., Morandi, A., Jouret, B., Landt, O., Tounian, P., Levy-Marchal, C., Buzzetti, R., Pinelli, L., Balkau, B., Horber, F., Bougnères, P., Froguel, P., Meyre, D. (2007) Nonsynonymous polymorphisms in melanocortin-4 receptor protect against obesity: The two facets of a Janus obesity gene. Hum. Mol. Genet. 16, 1837–1844.

Subramanian, A., Tamayo, P., Mootha, V.K., Mukherjee, S., Ebert, B.L., Gillette, M.A., Paulovich, A., Pomeroy, S.L., Golub, T.R., Lander, E.S., Mesirov, J.P. (2005) Gene set enrichment analysis: a knowledge-based approach for interpreting genome-wide expression profiles. Proc Natl Acad Sci U S A 102, 15545–15550.

Szustakowski, J.D., Balasubramanian, S., Sasson, A., Khalid, S., Bronson, P.G., Kvikstad, E., Wong, E., Liu, D., Davis, J.W., Haefliger, C., Loomis, A.K., Mikkilineni, R., Noh, H.J., Wadhawan, S., Bai, X., Hawes, A., Krasheninina, O., Ulloa, R., Lopez, A., Smith, E.N., Waring, J., Whelan, C.D., Tsai, E.A., Overton, J., Salerno, W., Jacob, H., Szalma, S., Runz, H., Hinkle, G., Nioi, P., Petrovski, S., Miller, M.R., Baras, A., Mitnaul, L., Reid, J.G. (2020) Advancing Human Genetics Research and Drug Discovery through Exome Sequencing of the UK Biobank. medRxiv 2020.11.02.20222232.

Thaker, V. V (2017) GENETIC AND EPIGENETIC CAUSES OF OBESITY. Adolesc. Med. State Art Rev. 28, 379–405.

Winkler, T.W., Justice, A.E., Graff, M., Barata, L., Feitosa, M.F., Chu, S., Czajkowski, J., Esko, T., Fall, T., Kilpeläinen, T.O., Lu, Y., Mägi, R., Mihailov, E., Pers, T.H., Rüeger, S., Teumer, A., Ehret, G.B., Ferreira, T., Heard-Costa, N.L., Karjalainen, J., Lagou, V., Mahajan, A., Neinast, M.D., Prokopenko, I., Simino, J., Teslovich, T.M., Jansen, R., Westra, H.J., White, C.C., Absher, D., Ahluwalia, T.S., Ahmad, S., Albrecht, E., Alves, A.C., Bragg-Gresham, J.L., de Craen, A.J.M., Bis, J.C., Bonnefond, A., Boucher, G., Cadby, G., Cheng, Y.C., Chiang, C.W.K., Delgado, G., Demirkan, A., Dueker, N., Eklund, N., Eiriksdottir, G., Eriksson, J., Feenstra, B., Fischer, K., Frau, F., Galesloot, T.E., Geller, F., Goel, A., Gorski, M., Grammer, T.B., Gustafsson, S., Haitjema, S., Hottenga, J.J., Huffman, J.E., Jackson, A.U., Jacobs, K.B., Johansson, Å., Kaakinen, M., Kleber, M.E., Lahti, J., Leach, I.M., Lehne, B., Liu, Y., Lo, K.S., Lorentzon, M., Luan, J., Madden, P.A.F., Mangino, M., McKnight, B., Medina-Gomez, C., Monda, K.L., Montasser, M.E., Müller, G., Müller-Nurasyid, M., Nolte, I.M., Panoutsopoulou, K., Pascoe, L., Paternoster, L., Rayner, N.W., Renström, F., Rizzi, F., Rose, L.M., Ryan, K.A., Salo, P., Sanna, S., Scharnagl, H., Shi, J., Smith, A.V., Southam, L., Stančáková, A., Steinthorsdottir, V., Strawbridge, R.J., Sung, Y.J., Tachmazidou, I., Tanaka, T., Thorleifsson, G., Trompet, S., Pervjakova, N., Tyrer, J.P., Vandenput, L., van der Laan, S.W., van der Velde, N., van Setten, J., van Vliet-Ostaptchouk, J. V., Verweij, N., Vlachopoulou, E., Waite, L.L., Wang, S.R., Wang, Z., Wild, S.H., Willenborg, C., Wilson, J.F., Wong, A., Yang, J., Yengo, L., Yerges-Armstrong, L.M., Yu, L., Zhang, W., Zhao, J.H., Andersson, E.A., Bakker, S.J.L., Baldassarre, D., Banasik, K., Barcella, M., Barlassina, C., Bellis, C., Benaglio, P., Blangero, J., Blüher, M., Bonnet, F., Bonnycastle, L.L., Boyd, H.A., Bruinenberg, M., Buchman, A.S., Campbell, H., Chen, Y.D.I., Chines, P.S., Claudi-Boehm, S., Cole, J., Collins, F.S., de Geus, E.J.C., de Groot, L.C.P.G.M., Dimitriou, M., Duan, J., Enroth, S., Eury, E., Farmaki, A.E., Forouhi, N.G., Friedrich, N., Gejman, P. V., Gigante, B., Glorioso, N., Go, A.S., Gottesman, O., Gräßler, J., Grallert, H., Grarup, N., Gu, Y.M., Broer, L., Ham, A.C., Hansen, T., Harris, T.B., Hartman, C.A., Hassinen, M., Hastie, N., Hattersley, A.T., Heath, A.C., Henders, A.K., Hernandez, D., Hillege, H., Holmen, O., Hovingh, K.G., Hui, J., Husemoen, L.L., Hutri-Kähönen, N., Hysi, P.G., Illig, T., De Jager, P.L., Jalilzadeh, S., Jørgensen, T., Jukema, J.W., Juonala, M., Kanoni, S., Karaleftheri, M., Khaw, K.T., Kinnunen, L., Kittner, S.J., Koenig, W., Kolcic, I., Kovacs, P., Krarup, N.T., Kratzer, W., Krüger, J., Kuh, D., Kumari, M., Kyriakou, T., Langenberg, C., Lannfelt, L., Lanzani, C., Lotay, V., Launer, L.J., Leander, K., Lindström, J., Linneberg, A., Liu, Y.P., Lobbens, S., Luben, R., Lyssenko, V., Männistö, S., Magnusson, P.K., McArdle, W.L., Menni, C., Merger, S., Milani, L., Montgomery, G.W., Morris, A.P., Narisu, N., Nelis, M., Ong, K.K., Palotie, A., Pérusse, L., Pichler, I., Pilia, M.G., Pouta, A., Rheinberger, M., Ribel-Madsen, R., Richards, M., Rice, K.M., Rice, T.K., Rivolta, C., Salomaa, V., Sanders, A.R., Sarzynski, M.A., Scholtens, S., Scott, R.A., Scott, W.R., Sebert, S., Sengupta, S., Sennblad, B., Seufferlein, T., Silveira, A., Slagboom, P.E., Smit, J.H., Sparsø, T.H., Stirrups, K., Stolk, R.P., Stringham, H.M., Swertz, M.A., Swift, A.J., Syvänen, A.C., Tan, S.T., Thorand, B., Tönjes, A., Tremblay, A., Tsafantakis, E., van der Most, P.J., Völker, U., Vohl, M.C., Vonk, J.M., Waldenberger, M., Walker, R.W., Wennauer, R., Widén, E., Willemsen, G., Wilsgaard, T., Wright, A.F., Zillikens, M.C., van Dijk, S.C., van Schoor, N.M., Asselbergs, F.W., de Bakker, P.I.W., Beckmann, J.S., Beilby, J., Bennett, D.A., Bergman, R.N., Bergmann, S., Böger, C.A., Boehm, B.O., Boerwinkle, E., Boomsma, D.I., Bornstein, S.R., Bottinger, E.P., Bouchard, C., Chambers, J.C., Chanock, S.J., Chasman, D.I., Cucca, F., Cusi, D., Dedoussis, G., Erdmann, J., Eriksson, J.G., Evans, D.A., de Faire, U., Farrall, M., Ferrucci, L., Ford, I., Franke, L., Franks, P.W., Froguel, P., Gansevoort, R.T., Gieger, C., Grönberg, H., Gudnason, V., Gyllensten, U., Hall, P., Hamsten, A., van der Harst, P., Hayward, C., Heliövaara, M., Hengstenberg, C., Hicks, A.A., Hingorani, A., Hofman, A., Hu, F., Huikuri, H. V., Hveem, K., James, A.L., Jordan, J.M., Jula, A., Kähönen, M., Kajantie, E., Kathiresan, S., Kiemeney, L.A.L.M., Kivimaki, M., Knekt, P.B., Koistinen, H.A., Kooner, J.S., Koskinen, S., Kuusisto, J., Maerz, W., Martin, N.G., Laakso, M., Lakka, T.A., Lehtimäki, T., Lettre, G., Levinson, D.F., Lind, L., Lokki, M.L., Mäntyselkä, P., Melbye, M., Metspalu, A., Mitchell, B.D., Moll, F.L., Murray, J.C., Musk, A.W., Nieminen, M.S., Njølstad, I., Ohlsson, C., Oldehinkel, A.J., Oostra, B.A., Palmer, L.J., Pankow, J.S., Pasterkamp, G., Pedersen, N.L., Pedersen, O., Penninx, B.W., Perola, M., Peters, A., Polašek, O., Pramstaller, P.P., Psaty, B.M., Qi, L., Quertermous, T., Raitakari, O.T., Rankinen, T., Rauramaa, R., Ridker, P.M., Rioux, J.D., Rivadeneira, F., Rotter, J.I., Rudan, I., den Ruijter, H.M., Saltevo, J., Sattar, N., Schunkert, H., Schwarz, P.E.H., Shuldiner, A.R., Sinisalo, J., Snieder, H., Sørensen, T.I.A., Spector, T.D., Staessen, J.A., Stefania, B., Thorsteinsdottir, U., Stumvoll, M., Tardif, J.C., Tremoli, E., Tuomilehto, J., Uitterlinden, A.G., Uusitupa, M., Verbeek, A.L.M., Vermeulen, S.H., Viikari, J.S., Vitart, V., Völzke, H., Vollenweider, P., Waeber, G., Walker, M., Wallaschofski, H., Wareham, N.J., Watkins, H., Zeggini, E., Chakravarti, A., Clegg, D.J., Cupples, L.A., Gordon-Larsen, P., Jaquish, C.E., Rao, D.C., Abecasis, G.R., Assimes, T.L., Barroso, I., Berndt, S.I., Boehnke, M., Deloukas, P., Fox, C.S., Groop, L.C., Hunter, D.J., Ingelsson, E., Kaplan, R.C., McCarthy, M.I., Mohlke, K.L., O’Connell, J.R., Schlessinger, D., Strachan, D.P., Stefansson, K., van Duijn, C.M., Hirschhorn, J.N., Lindgren, C.M., Heid, I.M., North, K.E., Borecki, I.B., Kutalik, Z., Loos, R.J.F. (2015) The Influence of Age and Sex on Genetic Associations with Adult Body Size and Shape: A Large-Scale Genome-Wide Interaction Study. PLoS Genet. 11, 1–42.

Xu, Y., Zhang, Q., Tan, L., Xie, X., Zhao, Y. (2019) The characteristics and biological significance of NPC2: Mutation and disease. Mutat. Res. − Rev. Mutat. Res.

Younis, S., Schönke, M., Massart, J., Hjortebjerg, R., Sundström, E., Gustafson, U., Björnholm, M., Krook, A., Frystyk, J., Zierath, J.R., Andersson, L. (2018) The ZBED6-IGF2 axis has a major effect on growth of skeletal muscle and internal organs in placental mammals. Proc. Natl. Acad. Sci. U. S. A. 115, E2048–E2057.

Zhang, X., Jiang, S., Mitok, K.A., Li, L., Attie, A.D., Martin, T.F.J. (2017) BAI AP3, a C2 domain-containing Munc 13 protein, controls the fate of dense-core vesicles in neuroendocrine cells. J. Cell Biol. 216, 2151–2166.

